# PINCS-1: protocol for a feasibility study investigating the acceptability and accuracy of cervical screening and self-sampling in women at 6-weeks postnatal

**DOI:** 10.1101/2024.11.20.24317620

**Authors:** Victoria Cullimore, Rebecca Newhouse, Holly Baker-Rand, Adam R Brentnall, Kim Chu, Karin Denton, Lorna McWilliams, Alex Sargent, Sudha Sundar, Emma J. Crosbie, Jo Morrison

**Author notes:** **Corresponding author** Dr Jo Morrison. Twitter handles @victoriacullim1; @DrJoMorrison1; @ProfEmmaCrosbie; @NewhouseRebecca; @sundar_sudha. Bluesky @jomorrison.bsky.social.

## Abstract

**Introduction:** Cervical screening rates in the UK are falling, limiting our ability to prevent cervical cancer. Peak incidence of cervical cancer coincides with average age of childbirth and women with young children are less likely to be screened. Current guidelines advise waiting 12-weeks after delivery to perform cervical screening, but this recommendation is not based on evidence from the era of liquid-based cytology (LBC) or high-risk Human Papilloma Virus (hrHPV) testing. New mums suggested that cervical screening could be offered at 6-weeks post-delivery, in conjunction with the postnatal check-up with the general practice team in primary care.

**Methods and analysis:** A study of 100 participants will be performed to assess feasibility and acceptability of cervical screening at 6- and 12-weeks postnatal, with urine self-sampling at each time point. This will inform whether women are prepared to undergo cervical screening at 6-weeks postnatal and feasibility of a future pair-wise diagnostic test accuracy study, or whether alternative study designs are needed. At each appointment, participants will complete a questionnaire about their experience and thoughts regarding screening. Sub-studies ask participants who withdraw or decline their reasons, to identify barriers. The study will move to a second phase, when 100 participants will be individually randomised to sampling at 6-weeks or 12-weeks, once 100 participants have completed the 6-week screen in PINCS-1, or recruitment is poor, indicating that a paired-sample design is not feasible.

**Ethics and dissemination:** Ethical approval for PINCS-1 was given by the Stanmore Research Ethics Committee. The results, including participant feedback at each stage, will inform design of large studies to determine accuracy and clinical impact of cervical screening at 6-weeks postnatal, identifying whether giving choice will improve screening uptake. Data will inform sample size needed for future studies to have adequate power. Results will also inform future NHS Cervical Screening Programme management. Results will be shared via scientific publication and via conventional and social media channels accessed by young women.

**Strengths and limitations of PINCS-1:** *Strengths:*

- The first study to focus on acceptability and reliability of screening, including self-sampling in postnatal women, to test hypothesis and generate data to inform further study design, following recommendations by Elridge et al.^1^
- Multiple points at which acceptability will be assessed by collecting participants’ views and participant-reported outcomes.
- Offering self-screening at the time of another appointment was a successful strategy in the YouScreen study.^2^

*Limitations:*

- This study has a limited sample size and is not statistically powered to evaluate the diagnostic test accuracy or the impact of offering screening during postnatal visits on overall screening uptake
- Screening will be performed in secondary care settings throughout this study. However, anticipated changes to screening would be expected to be relevant to primary care in the future studies.

## Introduction

Cervical cancer is one of the most preventable malignancies encountered worldwide, due to a combination of primary prevention (HPV vaccination) and secondary prevention (cervical screening) strategies. Global elimination of cervical cancer is a key World Health Organisation strategy.^3,4^ By 2022, cervical screening coverage rates in England had fallen to 66% in women/people with a cervix aged 25-49 years, and to less than 50% in some areas. This is markedly below the National Health Service Cervical Screening Programme (NHS CSP) standard of 80%. The majority of cervical cancers now occur in under-screened women ^5–7^. Women with young children under 5 years of age are less likely to participate in cervical screening, as are individuals from ethnic minority backgrounds and lower socioeconomic groups, and these groups are also more likely to have had more children and at a younger age.^8^

Peak incidence of cervical cancer in the UK between 2016 and 2019 was in the 30-34-year-old cohort, followed by cases in women aged 25 to 29.^9^ This coincides with the average age of mothers giving birth in England and Wales of 30.9 years.^10^ Our local cervical cancer audit between 2016 and 2017 identified that 15% of women diagnosed with cervical cancer were currently, or had recently been, pregnant and had been eligible for cervical screening in pregnancy or postnatally, but none had attended. We found that 50% of women were overdue for cervical screening by the end of their pregnancy and by 6 months postnatal more than half had still not attended screening.^11^ This quality improvement (QI) project included canvassed views of new mothers/parents and primary care providers, through focus groups, which identified causes of poor uptake and generated ideas for change.^11^ One idea, from both new mothers and primary care practice staff, was to offer postnatal cervical screening at the 6-week postnatal check-up, facilitating easier attendance for women by reducing barriers.^12^ Self-testing for high-risk Human Papilloma Virus (hrHPV) was also suggested to improve screening uptake. Interestingly, offering opportunistic self-screening at a GP appointment, was demonstrated to be an effective strategy in the recent YouScreen study, potentially leading to a 7.6% improvement in overall screening rates.^2,13^

There are numerous barriers to screening in young women, including a perception that this age group are not at risk, inadequate knowledge, and fear of pain, discomfort and embarrassment. However, being busy and not getting around to having a test were independent factors, regardless of screening status^14,15^ Our work showed that we could improve uptake by 8% in the postnatal cohort, largely by improving education of midwives and women in pregnancy.^11^ Detailed quantitative and qualitative feedback in the pre-PINCS acceptability study (unpublished data) alongside the previous QI project focus groups, told us that new parents have many competing priorities and often struggle to address their own health needs.

NICE guidelines recommend a 6-week postnatal check for mothers and babies, which is attended by 78% of eligible people.^16,17^ This appointment provides an opportunity for healthcare professionals to discuss multiple topics: infant feeding, lifestyle advice, contraception and health promotion, including discussion of cervical screening.^17,18^ New mothers and primary care staff told us that offering to combine this visit with postnatal cervical screening would remove a significant barrier, particularly as ‘just putting it off’ was the most common reason for younger women being out-of-date for screening in a study by Jo’s Cervical Cancer Trust.^19^

National guidance currently advises waiting 12 weeks after childbirth for a routine cervical screening test if it was due in pregnancy.^20^ This recommendation is based on one comparison of conventional cytology with Papanicolaou smear testing at 4- vs. 6- vs. 8-weeks postnatal in just 55 participants.^21^ There were increased inflammatory changes in Papanicolaou smears taken earlier, leading to more false-positive, low-grade results. However, this pre-dates hrHPV primary testing (or triage) and liquid-based cytology (LBC), which dramatically improve the ability to test even inflammatory samples, and those contaminated by blood and lochia.

An Irish observational study, including 556 postnatal women, reported no difference in inadequate cervical sample rates when the cervical sample was taken at 6-weeks postnatal using LBC compared to a non-pregnant gynaecological population consisting of 1429 women.^22^ Using LBC appears to negate the previously held belief that postnatal cervical samples should be delayed until 12-weeks postpartum. HPV-testing was not conducted in this study and there have been no studies directly comparing LBC cervical screening samples at different postnatal time points in a diagnostic test accuracy (DTA) context. Furthermore, hrHPV infection rates are similar during and outside of pregnancy, although these studies performed hrHPV tests at varying postnatal intervals, ranging from 45 days ^23^ to 6-months^24^ and used swabs rather than clinician-collected LBC samples. This variation limits the applicability of these findings to current UK practice. The current recommendations to delay cervical screening until 12-weeks postpartum are therefore based on long-held perceived wisdom, rather than sound evidence of differences in DTA using current screening methods.

Many women struggle to undergo conventional cervical screening, especially those in higher-risk and socioeconomically disadvantaged groups.^25^ hrHPV testing using self-sampling methods offers an alternative and improves screening uptake in under-screened women.^26^ However, previous studies have not specifically targeted postnatal women, whose feelings on vaginal sampling may be affected by recent birth experiences. Our project also provides an opportunity to test the feasibility & acceptability^12^ of self-testing for hrHPV in urine samples at 6- and 12-weeks postnatal, alongside conventional testing.

We have investigated the acceptability of cervical screening earlier in the postnatal period in a quantitative and qualitative attitudes study (Pre-PINCS – National Institute for Health and Care Research (NIHR) Central Portfolio Management System (CPMS) ID: 55489). Preliminary analyses suggest that over two-thirds of respondents would be willing to take part in a clinical study of 6-week clinician-taken cervical screening and nearly 8 out of every 10 would be willing to take part in a study of self-testing with urine samples (unpublished results; n = 454). Over half of the participants agreed or strongly agreed that they would be more likely to have cervical screening if offered at the time of their postnatal check-up, with only 1 in 13 disagreeing or strongly disagreeing to this (unpublished results).

### Aim

PINCS is a two-phase study with a paired-sample study design (PINCS-1) performed at 6 and 12-weeks postpartum, followed by a randomised two-arm feasibility study in phase 2 (PINCS-2, which will be published as a separate protocol), comparing sampling at 6- or 12-weeks postnatal with the overall aim of assessing the acceptability and feasibility of these study designs in comparing cervical screening and self-testing at 6- versus 12-weeks postnatal.

### Objectives of PINCS-1

The primary objective is to evaluate a paired-sample study design investigating the acceptability of cervical screening at 6-weeks postpartum, willingness to have repeat screening at 12-weeks postpartum, and to evaluate the feasibility for a larger-scale trial.

The secondary objectives are:

1. To evaluate acceptability of clinician-taken cervical samples and self-collected urine samples for screening tests in those who decline, and in those who consent both at 6- and 12-weeks using questionnaire data.
2. To assess the quality of cervical samples from clinician-taken samples at 6-weeks postnatal.
3. To determine the agreement in hrHPV status at 6- and 12-weeks postnatal between clinician-taken cervical samples and self-collected urine samples.

## Methods and analysis

### Study design

PINCS-1 is a paired feasibility study to investigate the acceptability of cervical screening and urine self-sampling in postnatal women at 6-weeks and 12-weeks postnatal.

### Study setting

The primary study site will be Somerset NHS Foundation Trust. Several study sites across South West England will also collaborate in this study, recruiting participants, completing study visits and data collection. Somerset NHS Foundation Trust act as the study sponsor.

### Patient and public involvement (PPI)

This study was instigated following the direct request by stakeholders, when investigating methods to reduce barriers to cervical screening in recently pregnant women/people.^11^ Multiple ideas for change were generated through stakeholder groups involving new mothers, young women who had a cervical cancer diagnosed shortly after pregnancy, and primary care staff directly involved in both postnatal care and cervical screening. In addition to suggestions about improving education about cervical screening for midwives and pregnant women/new parents, both public and healthcare participants identified two areas to target: earlier postnatal screening potentially at the time of the postnatal GP appointment and the use of self-screening methods.

We worked with local Maternity Voices groups, whose members included women from marginalised communities, to design study materials, questionnaires and semi-structured interviews for the pre-PINCS study, which is currently undergoing analysis. Pre-PINCS was a two-phase study consisting of a questionnaire and in-depth qualitative analysis of semi-structured interviews. This was performed to gather information, from pregnant women and people within 5-years of their last childbirth, about the acceptability and feasibility of the PINCS studies; these results directly informed the PINCS study design and materials, with specific feedback from participants.

### Participants and recruitment

Potential participants will be identified by members of their existing clinical care team including GPs, community or hospital midwifes, health visitors, practice nurses or obstetricians, alongside the local research teams, both antenatally and up to 6-weeks postnatal. Potential participants may also self-identify through publicity literature on recruitment sites and via the social media channels of gynaecological cancer charities (e.g. GO Girls, Eve Appeal) and local and national social media groups for new mothers (e.g. Mumsnet). Publicity will be in the form of posters and leaflets, distributed via social media, at antenatal events, and at routine appointments or shared through the electronic maternity care record. Potential participants will be given a participant information leaflet and, if interested in taking part, they will be referred to a member of the study team. A screening and eligibility questionnaire will be completed with all potential participants and, if eligible and consenting to proceed, an electronic consent form will be completed with an investigator. Participants will be informed of their right to rescind consent at any point during the study and provided with information on how to do this.

Recruitment to PINCS-1 will end when at least 100 recruited participants have attended and completed both clinician-taken cervical sample and urine self-sample at the 6-week appointment and have attended or declined to attend their 12-week appointment. If participants withdraw prior to the 6-week sample, further participants will be recruited. In the instance of low recruitment, an earlier end point may be initiated following discussion with the Independent Trial Steering Committee (ITSC). Commencement of PINCS-2 will proceed after review of results of PINCS-1 by the ITSC to confirm that differences in testing at 6-week vs. 12-weeks are within acceptable limits to proceed safely.

#### Inclusion criteria

- 24.5 years (24 years and 183 days or greater on day of consent) to <65 years old
- Female with a cervix (regardless of gender identity)
- Currently pregnant or within 6-weeks of delivery
- Able to give informed consent

#### Exclusion criteria

- Absence of a cervix
- Not eligible for the NHS CSP
- Unable to give fully informed consent

The study is open to all those eligible for cervical screening, regardless of screening status. To understand the reasons for non-participation and to establish an uptake rate, a cohort of 100 potential participants will be approached and the acceptance rate recorded. All those who decline to participate will be given the opportunity to describe the reasons behind this. All participants who initially consent to the study, but choose to withdraw, will be offered a short electronic questionnaire to identify any concerns and barriers to participation.

### Sample size

This study will aim to recruit at least 100 participants to PINCS-1. This sample size was chosen following findings from the pre-PINCS study regarding manageable recruitment in postnatal patients as well as input from statisticians and other experienced researchers with experience in feasibility studies. PINCS-2 will aim to recruit another 100 participants, randomised to either 6- or 12-week testing, with self-sampling with both urine and vaginal swabs at the same visit, allowing direct comparison of acceptability in this cohort. This sample size will provide a standard error on uptake at most 2.5% on each proportion, which we judge to be suitable for assessing acceptability and feasibility of a subsequent paired study design for accuracy. It will inform us as to how prepared women are to undergo cervical screening with a speculum examination at 6-weeks postnatal, and the feasibility of a paired-sample design using repeat testing in the same participant with clinician- and/or self-samples at both, or either, time points.

### Study visits

The study will consist of a screening and consent appointment followed by two study visits (see Figure 1). At each study visit, participants will undergo clinician-taken cervical screening samples using a speculum examination and cervical sample/sweep test, for hrHPV testing and cytology at 6-weeks postnatal. They will also undergo hrHPV testing using urine samples collected in a Colli-pee® device at both time points, to ascertain the agreement with clinician-taken sampling and the acceptability to participants at both time points.^27^

**Figure 1:**
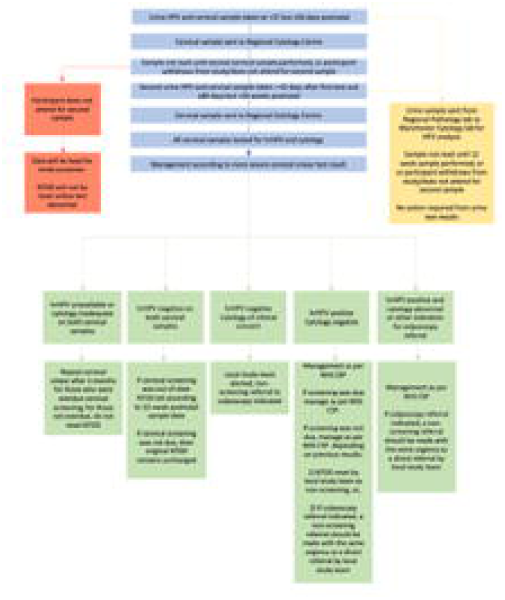
PINCS-1 participant flowchart. NTDD = Next Test Due Date.

We will perform a patient questionnaire after sampling (web-based or paper), at both 6- and 12-weeks, to ascertain acceptability (concordance with protocol), feasibility (ability to recruit), patient-reported outcomes, including discomfort of testing, preferences regarding timing of screening and attitudes to introducing the option of screening at the 6-week postnatal check up in the GP practice.

### Management of cytology and urine samples

Cytology samples performed following a hrHPV positive test will be dual labelled with patient identifying information and study details/study number and stored and managed in accordance with NHS CSP guidance.

Results of the cytological assessment on hrHPV negative samples, which would not ordinarily be performed as part of the NHS CSP, will not be uploaded to the NHS Cervical Screening Administration Service (CSAS), but will be recorded for the purposes of the study and acted on within the study protocol. Cytology samples from hrHPV-negative tests at 6-weeks postnatal will be destroyed at the end of the study period and not made available to CSAS for future audit.

Management of results and further cervical screening will depend upon previous cervical screening history (whether up to date at time of study, or not), attendance for both samples, and results of screening (see Figure 2 and Figure 3). Participants will be contacted with results and management plan, questions about further management answered, and asked about any adverse events, as well as being encouraged to self-report adverse events to the study team.

**Figure 2:**
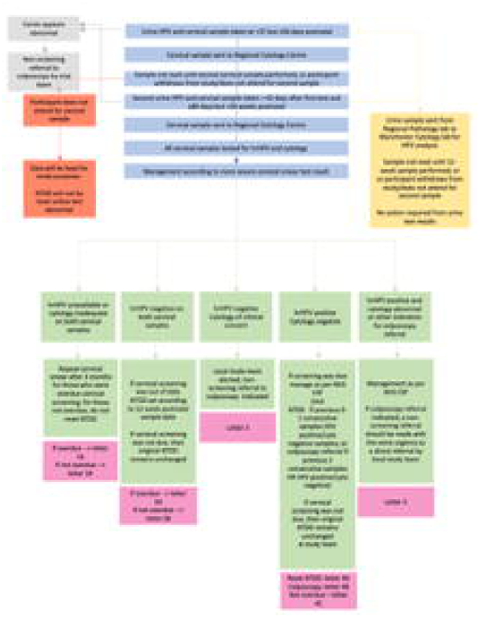
PINCS-1 study flowchart for those having samples at 6- and 12-weeks. NHS CSP = NHS Cervical Screening Programme; NTDD = Next Test Due Date; hrHPV = high risk Human Papilloma Virus; DTA = diagnostic test accuracy; PROM = patient reported outcome measures

**Figure 3:**
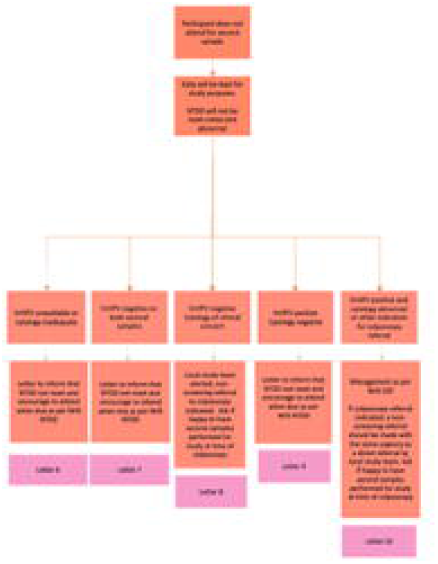
PINCS-1 study flowchart for those having samples at 6-weeks who do not attend for their 12-week sample. NHS CSP = NHS Cervical Screening Programme; NTDD = Next Test Due Date; hrHPV = high risk Human Papilloma Virus.

Urine samples will be labelled with the study details and study ID number and will be destroyed after testing and communication of results with the study team.

### Data collection

Each participant will be assigned a unique study ID following consent to participate. All trial data will be uploaded to the secure web application for managing data, REDCap, which will host the electronic Case Report Form (eCRF). The study co-ordinators will be responsible for analysing and monitoring the data from all sites and thus will have full access to the inputted information and local investigators will be able to access the data from their site only.

### Statistical analysis

Full details of the statistical analysis will be described in a statistical analysis plan that will be written and finalised before data lock. The primary outcomes are binary variables. We will estimate 95% CIs for each using Wilson’s method.

## Discussion

Enhancing cervical screening uptake is a healthcare priority, as adequate screening rates lead to reduced incidences of precancerous and cancerous changes in the cervix.^6,7^ There is a clear need for research in methods to improve attendance of cervical screening in younger women due to a lack of proven strategies in the current literature.^28^ Pregnancy provides several points of contact to engage patients in health promotion through the increased access to healthcare and provides a valuable opportunity to educate and organise cervical screening, especially in ‘hardly reached’ groups.^10,11,29^ Offering opportunistic self-sampling in a healthcare setting during a pre-existing appointment with vaginal swabs to non-attenders achieved uptake rates of 55.9% in a recent study, compared with only 12.9% of those sent test kits via direct-mail.^2^ They found that urine self-sampling was preferred to vaginal sampling (41.9% vs. 15.4%), especially among women from ethnic minorities.^13^ From our preliminary unpublished attitudes to self-sampling data, this is likely to be even more pertinent to the postnatal cohort. However, this work also highlighted that the idea of self-sampling is not preferable to all. The data from YouScreen support our hypothesis that offering increased choice, and opportunities for testing when people are otherwise attending primary care appointments, is important to improve screening rates. Women have identified making and attending appointments as a significant barrier to screening and therefore it is essential to minimise process-based restrictions that limit accessibility to screening services.^15,30^ Combining screening with postnatal check-ups offers a golden opportunity to inform women, promote self-care and provide low-effort access to screening. This may require increased flexibility of primary care appointments, unless self-sampling is accurate enough to allow this as an alternative and support a redirection to focus of postnatal care on maternal healthcare needs, not just those of their babies.

We outline the protocol for a study evaluating the feasibility and acceptability of cervical screening using pair-wise sampling of clinician-taken cervical screening tests and self-testing with urine samples at 6- and 12-weeks postnatal. Providing there is minimal difference in inadequacy rates of screening and hrHPV positive rates at 6- and 12-weeks in PINCS-1, which is not anticipated based on previous data, we will perform a second feasibility study (PINCS-2) that aims to recruit 100 participants who will be randomised to LBC screening at 6- or 12-weeks postnatal. Urine and vaginal swab self-sampling will be performed at the time of LBC screening. This further study will assess feasibility of individual consent and randomisation. Uptake to the study, and acceptability of LBC screening at 6-weeks in the consented study sample, will inform whether progression to a definitive trial is justified N=50 participants per arm will provide precision of at least 3.5% on the proportion who accept the invitation, which we judge sufficient to determine feasibility. A major amendment to our ethics agreement will be required for PINCS-2 and a separate open protocol will be published once this is in place. For both feasibility study phases (PINCS-1 and PINCS-2) we will invite women to join regardless of screening status at the end of pregnancy, to maximise participation. We will conduct subgroup analyses of uptake by screening status to determine feasibility of then doing the same for the definitive study.

Overall, through the PINCS studies we anticipate establishing the level of acceptability and feasibility to inform design of two further studies and which is best to take forward. First, a DTA study to determine the accuracy of screening for hrHPV and cytological abnormalities at 6-weeks postnatal. This will compare the inadequacy rates, sensitivity and specificity of cervical screening at 6- versus 12-weeks postnatal, informing whether offering earlier postnatal screening is accurate. Provisional power calculations, based on inadequacy rates, estimated requiring over 1000 participants for a formal DTA of cervical screening at 6-week postnatal, hence why this feasibility study is required before embarking on such a significant undertaking. Data from PINCS-1 will inform this study design and size for adequate power.

Second, a randomised control trial (RCT) to examine the effect of earlier postnatal screening on screening uptake rates, as well as the longer-term clinical outcomes, such as rates of high-grade cervical intraepithelial neoplasia (CIN) at subsequent screening tests. Our proposed feasibility studies will determine whether, in this future RCT, it is reasonable and cost-effective to randomise individual participants to screening at 6- or 12-weeks. If this design is not feasible, a different design will be needed. For example, randomisation without prior consent, such as through applying for a CAG-251 exemption, or a pragmatic cluster-randomised design, such as that employed with YouScreen.^2^

Self-administered vaginal swabs and urine samples for hrHPV testing are under-evaluation.^2,27,31^ However, this research will provide crucial insights into postnatal individuals’ experiences with, and preferences for different self-sampling methods. These data will help determine the appropriate sample sizes needed to evaluate the accuracy and safety of these self-sampling techniques in future studies involving postnatal cohorts, as well as and influencing future changes to the NHS CSP.

## Ethics and dissemination

### Ethics

Ethical approval for PINCS-1 was granted by the Stanmore Research Ethics Committee for this study (IRAS project ID:321696; REC reference:24/LO/0206), was adopted by the NIHR Clinical Research Network (CRN) Portfolio (CPMS ID 60494) and is registered on the International Standard Randomised Controlled Trial Number (ISRCTN) registry (ISRCTN10071810; https://doi.org/10.1186/ISRCTN10071810).

### Publication and dissemination plan

Study results will be published as a PhD thesis and high impact peer-reviewed papers, as well as presentations at national and international meetings. They will also be presented to Maternity Voice Groups, gynaecological oncological charities, Mumsnet and local maternity social media sites. Any data arising from this study will be published and presented in an open-access peer-review journal. The manuscript will be deposited with the University of Exeter, according to the University of Exeter’s policies and data sharing policies.

### Individual participant data sharing statement

To ensure participant anonymity is safeguarded and subject to any reasonable and necessary delay, pseudonymised research data will be securely archived to a repository following publication of the results where they will be stored indefinitely. These data may be used in future research, here or abroad, and shared, subject to reasonable requests, approved by the sponsor, host institution and the regulatory authorities.

## Data Availability

All data produced in the present study are available upon reasonable request to the authors

## Acknowledgements

We are grateful to Flora Darch, Karen Tanner, Laura Courtney-Stamp, Tessa Dean and Richard Innes of the Department of Research and Development at Somerset NHS Foundation Trust; Kath Hunt, Andrzej Karmolinski and Nichole Villeneuve from the North Bristol NHS Trust for invaluable support and advice; and the NHS CSP Research Innovation and Development Advisory Committee for their formal and informal feedback, helpful suggestions and support in setting up this study. We also thank the public and patients involved in our previous QI and qualitative work that instigated and helped design this research.

## Authors’ contributions

All authors contributed to the study conception and design. The first draft of the manuscript was written by VC and revised by JM, HB-R, KC and RN. All authors have approved the final version and JM acts as guarantor.

## Protocol version

Version 2.8 – date 8/8/24

## Trial Registry

ISRCTN10071810 https://doi.org/10.1186/ISRCTN10071810

### Secondary identifying numbers

CPMS 60494, MR/X030776/1, IRAS 321696

## Sponsor information

### Organisation

Somerset Partnership NHS Foundation Trust

### Sponsor details

Musgrove Park Hospital Taunton

TA1 5DA

England

United Kingdom

+44 (0)1823333444

## Conflicting interests statement

VC - none to declare

RN - none to declare

HBR - none to declare

ARB – none to declare

KC – received honorarium and support from SeeGene

KD – received expenses and honorarium from Hologic. Received test kits and consumables from Hologic, Roche, Rovers and Copan for a previous study, now submitted for publication.

LMcW – none to declare

AS - A member of various expert groups providing advice to the English Cervical Screening Programme including on HPV self-sampling; holds an honorary contract with the University of Manchester to support research into HPV testing in urine samples and Professional Clinical Advisor to the English Cervical Screening Programme.

SS - none to declare

EJC - none to declare

JM – Clinical Advisor to the NHS Cervical Screening Programme Research Innovation and Development Advisory Committee.

